# VACtrac: Enhancing access immunization registry data for population outreach using Bulk Fast Interoperable Healthcare Resource (FHIR) protocol

**DOI:** 10.1101/2022.11.12.22282232

**Authors:** Leslie Lenert, Jeff Jacobs, James Agnew, Wei Ding, Katie Kirchoff, Duncan Weatherston, Kenneth Deans

## Abstract

COVID-19 vaccination uptake has been suboptimal, even in high-risk populations. This paper describes work to extend the Bulk Fast Healthcare Interoperability Resource (FHIR) standard for use in querying state Immunization Information Systems (IIS). We also describe a population vaccination outreach tool that uses both Bulk FHIR and automated single queries to access IIS data. Bulk FHIR protocols needed to be extended to support IIS’s responses for care outside an institution resulting in the addition of Group and Master Data Management FHIR profile functionalities to Bulk FHIR queries to support more accurate and easier retrieval of data. While real-world testing of Bulk FHIR queries using the vaccination outreach system was not possible, we tested an automated-single-query tool in a focused effort to reach 1500 high-risk patients. Results confirmed the potential for performance problems during periods of high demand that could be resolved by Bulk FHIR’s asynchronous retrieval methods.

## Introduction

COVID-19 pandemic has made the long-standing clinical problem of managing vaccine administration in target populations more apparent. Knowing who has and has not been “fully” vaccinated in a designated clinical population has always been difficult, especially when there are long gaps in care and vaccination requirements are continually changing. Further complicating the issue is the administration of vaccines in multiple sites by different clinical providers including pharmacies that are not connected electronically to population health care providers.[1] Moreover, the problem is not limited to COVID-19. Lack of access to in-person care has also created gaps in other routinely recommended vaccinations, even in previously well-managed populations.[2,3]

To help providers know which patients have and have not received age-appropriate vaccinations, public health jurisdictions have created Immunization Information Systems (IIS) that link and store immunization data across providers in their jurisdiction.[4] IIS are an effective intervention to improve vaccination uptake and help close vaccination gaps. [5] These systems are often driven by automatic feeds of data from electronic health records systems required under Meaningful Use regulations.[6]

At the individual patient level, in many EHR systems, it is relatively easy to query a state IIS system to check the vaccination status of a specific patient through existing application programming interfaces (APIs) of IIS systems. [7] Population health management is more complex. Routinely checking large lists of patients, one at a time, using existing protocols, for changes in vaccination status, can create functional challenges for IISs, clinical staff and providers. IIS interfaces to EHRs work well for accessing data onindividual patients, but typically do not allow for bulk review of an entire patient panel, for example. The extra work of reviewing patients one-at-a-time can negatively impact timeliness of care and efficiency of clinical staff. From a technical perspective, the number of queries for population health practice (potentially all patients, every day) is far higher than for clinical practice (patients visiting a provider for primary care or other services where vaccinations may be administered)[8]. A recent monograph estimated that COVID-19-based population health activities increased the load on state IIS by as much as 10-fold.[9]

One possible solution to the problem, advocated by Lenert and colleagues [1] is the enhancement of state IISs with Bulk Fast Healthcare Interoperability Resource (FHIR) [10,11] compatible systems, where providers can submit lists of patients for queries and receive back results asynchronously, to improve the logistics of response. To date, bulk FHIR query and response, also called Flat FHIR in FHIR Bulk Data Access Implementation Guide, capabilities have not been deployed or tested in the vaccine space in an operational IIS. This paper describes an implementation of prototype based on this model including:

- A Fast Healthcare Interoperability Resource standard database that mirrors Immunization Information system content and makes this content available by the Bulk FHIR standards to enhance access to IIS data by providers.
- A population health management program for use by providers and affiliated community organizations that can maintain a list of patients for a provider-based population-based outreach effort and communicate with a state IIS to maintain up-to-date records on the vaccination status of that population using either the Bulk FHIR protocol or repeated one-at-a-time queries..

These two components come together to form a system called VACtrac, which is a system designed to both enhance IIS bulk response capabilities and to support providers’ use of these capabilities in population health management. We implemented both components and used this testbed to explore extensions needed to FHIR protocols for Bulk queries of vaccine data and to understand how population health applications need to work in concert with Bulk FHIR enhanced IIS both in small and expanded scale studies.

## Methods

Figure 1 depicts the FHIR components of the VACtrac system. This system combines a FHIR immunization database that responds to bulk FHIR queries, connected to an IIS (left most box) and a population health management program that can query the FHIR database using either bulk or one-at-a-time querying methods (middle box).

**Figure 1.**
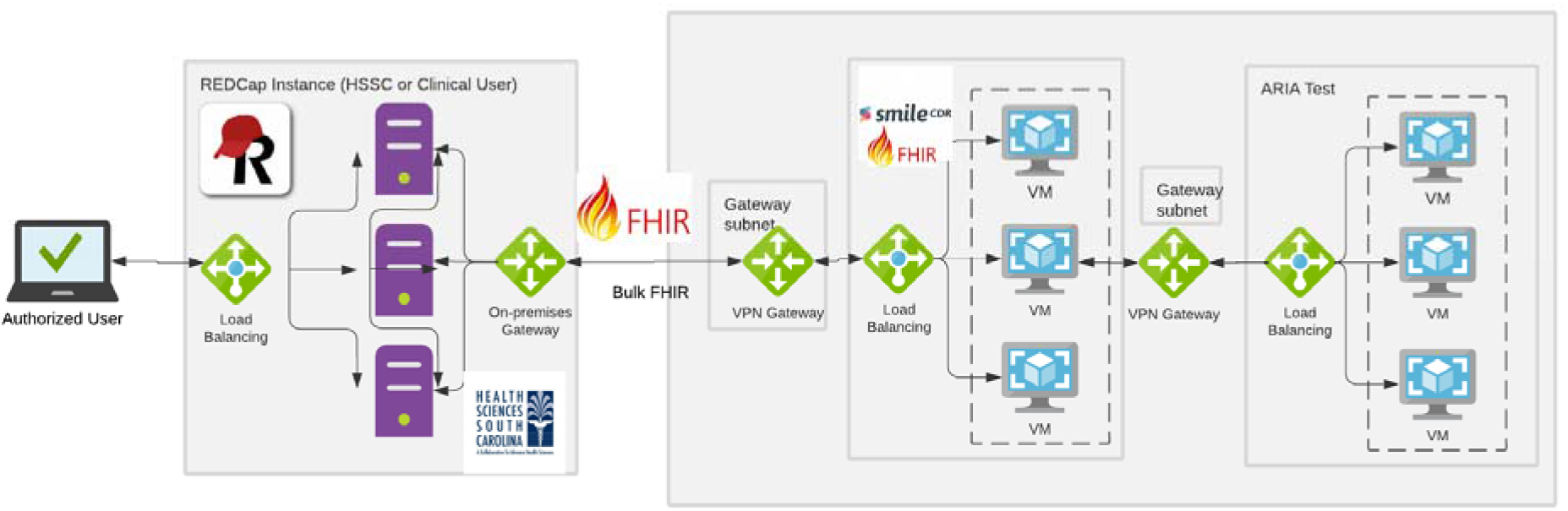
Architecture of VACtrac vaccination tracking tool that combined a FHIR server that replicates IIS vaccination data and a REDCap-based population outreach tool.

A significant focus of work was the design of a bulk FHIR server to respond to immunization queries. We achieved this by replication of the data in an IIS in the HAPI standard FHIR server (Smile CDR, Toronto, CN). Replication of the data streams in an IIS system in a FHIR repository was achieved by both the duplication of a stream of Health Level Seven International (HL7) V2.5 vaccination messages coming into the IIS from “connected” EHRs, and by periodic export of the processed data in the IIS to the FHIR database with synchronization of data at a patient level. We believed replication of HL7 v2.X vaccination data streams might work better where a single stand-alone solution was required to enhance access to results or in conjunction with a Health Information Exchange. Replication of an IIS’s database, in a FHIR server in a public health department FHIR server, might be a better option where state laws require public health control of IIS data and access. We implemented both approaches as part of testing. The replicated instance of VACtrac was designed to be used in a hybrid-cloud architecture, with the FHIR server residing in a cloud setting, to allow rapid scaling of computing resources to meet demand.

The specific use case we tested with VACtrac was monitoring of COVID-19 vaccination status of groups of patients, tracking the types of COVID-19 vaccines administered, the number of doses and the dates of vaccination, and integrating this data into the population health management portion of the program. Two sub-use cases were envisioned:

⍰ Use case (1) access of the FHIR server by a healthcare provider that may (1a) or may not (1b) have previously submitted vaccination data to the IIS along with demographic data in the message to identify patients needing follow-up for vaccination or submitted only a partial vaccination record.
⍰ Use case (2) access of the FHIR server by a school or university or use by an insurer to check the vaccination status of persons without a prior history of submission of vaccination record.

To implement these use case, we used an agile development approach, testing bulk queries against the FHIR database. Based on the responses, we identified extensions necessary to improve the precision and completeness of responses. We worked with the authors of the HAPI FHIR server (JA, DW) and implemented extensions to the FHIR environment and deployed a new instance of the modified FHIR server in the American Immunization Registry Association sandbox environment for IIS systems, further testing the ability of the FHIR server to ingest large numbers of HL7 V2.X test messages and respond to bulk queries of individuals.

### Population health management platform

The population health management platform was built around a low-cost (free to academic users), highly accessible database platform that offers a secure and Health Insurance Portability and Accountability Act-compliant platform (Research Electronic Data Capture (REDCap)) and was designed to be deployed in resource-challenged settings with volunteer staff from schools, churches, or other community organizations using a web interface. VACtrac supports providers with less advanced electronic health records systems that do not have population health management functionality, allowing users to upload spreadsheets with patient demographic, contact, and health data. Alternatively, VACtrac can extract clinical data for its list of patients via scheduled query of the Medical University of South Carolina’s (MUSC) research data warehouse or using REDCap’s Clinical Data Interoperability Services (CDIS) FHIR plug-in module for the Epic and Cerner EHRs. [12]

The REDCap database was designed to be used simultaneously by multiple providers, distributing the task of follow-up of patients across groups of case managers, with each group assigned a subset of patients for coordinated outreach. It limited access for a case manager to his or her designated set of patients at a time to preserve patients’ privacy. The RECDap instance also managed clinical and non-clinical data on individual patients’ risk that was used to prioritize outreach. The risk was conceptualized in two ways: using social determinants of health associated with patients’ residential neighborhoods (census tract) and based on clinical risk factors for the progression of COVID-19 infection to severe disease. Figure 2 shows example screens from the REDCap application.

**Figure 2.**
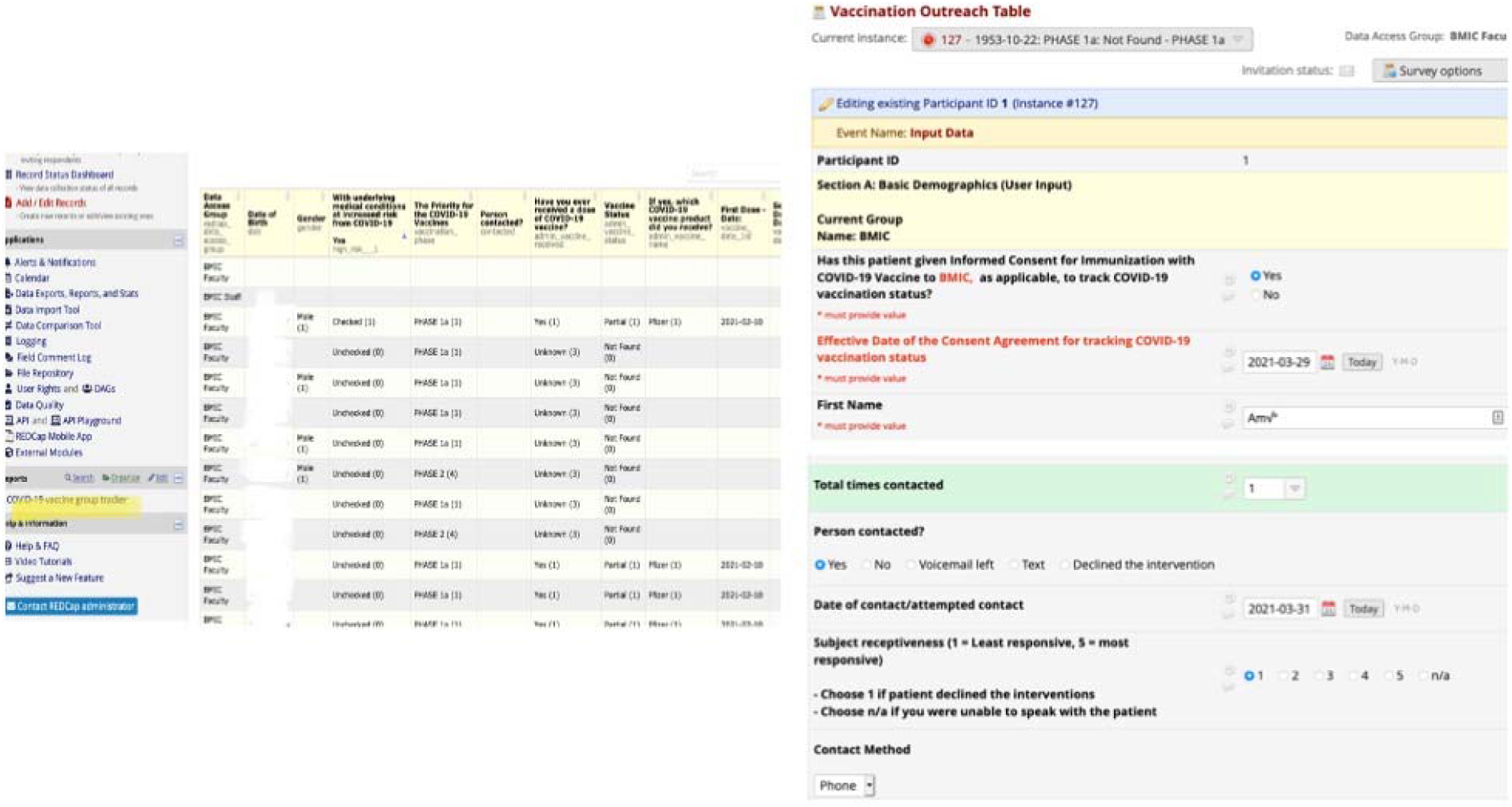
REDCap population health management application

REDCap software used custom middleware extensions to support queries of the vaccine database, allowing us to test our proposed extensions to the bulk FHIR protocol. To support access to IIS systems that did not have bulk FHIR capabilities, the REDCap instance also included software to cycle through its list of patients, one patient at a time, on a daily basis. The data flow of this application is shown in Figure 3. The application checked the IIS database using an HTTP SOAP query, with an automated login and automated cycling over its patient list. It used a variety of queues to manage waits and failures in the updating process, as shown in the figure.

**Figure 3.**
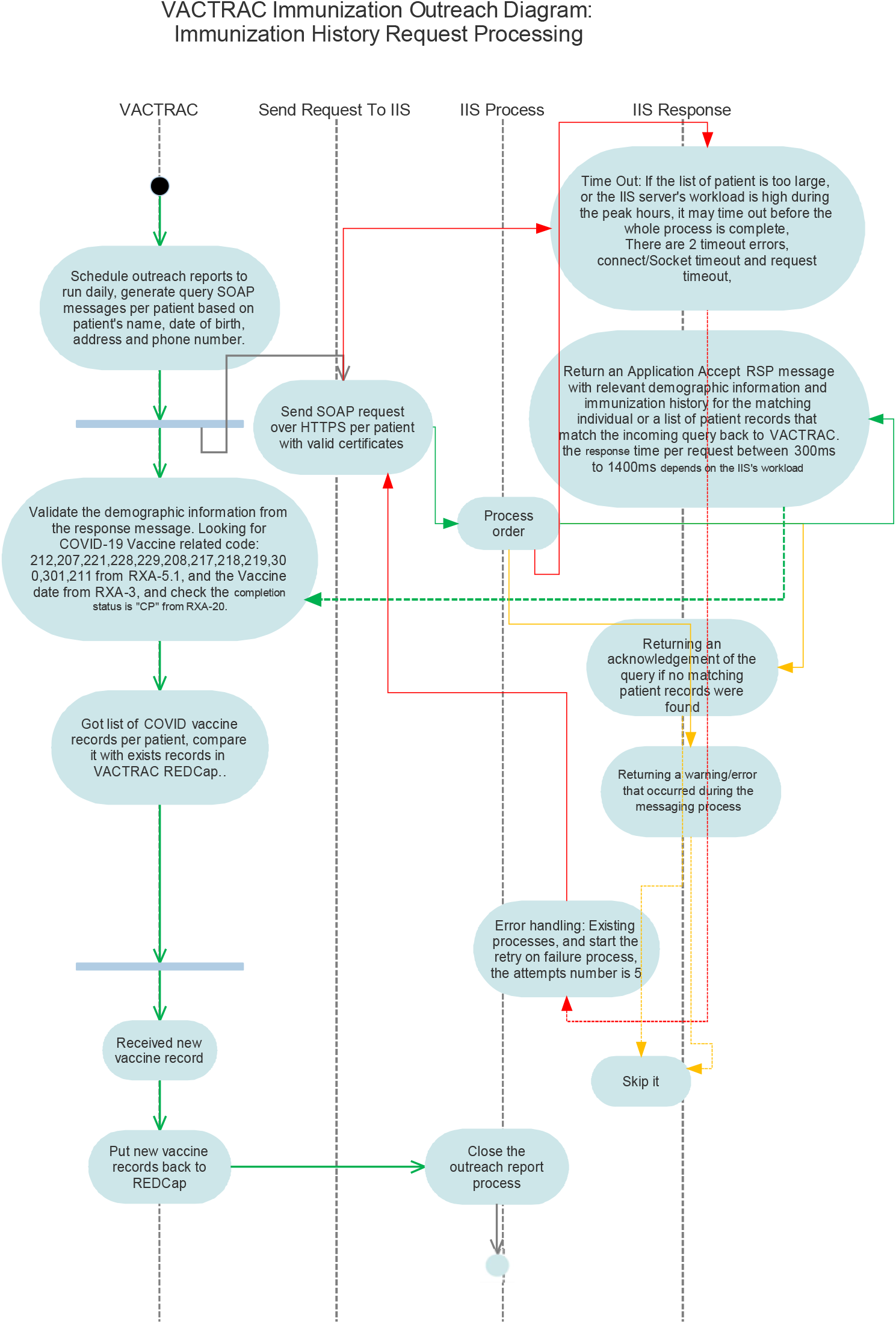
Data flow for one-by-one query from REDCap

### Population Health Evaluation Study

We tested the components of the VACtrac system in a quality improvement study comparing different approaches for prioritizing outreach to patients. One group focused on patients residing in high social need areas. Patients’ addresses were used to assign residences at the block level based on Federal Information Processing System (FIPS) codes. FIPS codes were then mapped to the JVION Vaccination Prioritization Index. [13] Patients who had undergone prior care at the Medical University of South Carolina since 1/1/2018, who had no prior record of COVID-19 vaccination in South Carolina’s SIMON registry, were randomly selected for inclusion based on and assigned to one of two groups:

Group 1: residence in high, very high or highest risk areas based on the JVION index, age greater than 18 years, and no EHR-documented COVID-19 vaccine within the MUSC Health System.

Group 2: residence in very low, low, or medium risk areas based on the JVION Vaccination Prioritization Index AND one or more CDC COVID Risk factor chronic conditions [14] AND no EHR-documented COVID-19 vaccine within the MUSC Health System

Patients were assigned to case managers by FIPS code area (so that resources could be coordinated) in blocks of 15 persons at a time, focusing on the most disadvantaged or highest illness level in each experimental group. Outreach was conducted by telephone, email, and text messaging, as supported by the REDCap application.

The South Carolina State Department of Health and Environmental Control chose not to participate in a replication of its SIMON database using a FHIR server. As a result, we used the patient-by-patient query mechanism for SIMON in our pilot study. This mechanism had good performance in testing in our internal American Immunization Registry Association (AIRA) sandboxes for updating of patients’ vaccination status in groups of hundreds of patients. Medical University of South Carolina student volunteers conducted a telephone outreach effort for COVID-19 vaccination using the REDcap application. Results of their outreach efforts were recorded in the database, including successful contact and intent to seek vaccination. Performance of the one-by-one algorithm was quantified using log files of the app to identify different potential failure types (REDCap app timing out the server query, IIS timing out REDCap session, and IIS failing to return patient data.)

The over study was classified as a quality improvement activity by the MUSC IRB.

## Results

### Adaptations required to the HAPI Standard FHIR Server

Initial work implementing Bulk FHIR Queries revealed that the requirement for transmitting a list of patient identifiers with query was awkward when applied to repetitive queries of thousands (or hundreds of thousands) of persons. When providing a list of identifiers, the implementation of the FHIR server may pass that list to the database server. There are limits to the number of parameters passed in a database query. There are also limits to the total size of a single query string. The exact sizes are determined by the underlying database server.

In FHIR, the Group resource is defined as a collection of entities and lends itself well to managing the set, or group, of patients in FHIR. Instead of locally managing the group of patients and passing the set of identifiers in GET or POST operations, the group is managed in FHIR whereby just the single group identifier is passed. For example, the following sample query would return the identified Pfizer COVID vaccine for all patients within group 123:

*Immunization?vaccineCode=http://hl7.org/fhir/sid/cvx|300&_has:Patient:subject:_has:Group:member:_id=Group/123*

In use case 1a, the querying system submits a list of patients for which it has previously submitted vaccination data along with demographic data. However, we assume that the Bulk FHIR querying system does not have access to the IIS system master person identifier or unique MDM FHIR identifier. Even so, Bulk FHIR queries for data in the IIS needed to be expanded beyond the direct match with a submitted institutional identifier, lest the system only return the results for the querying institution’s own data. To overcome this, an additional step and new capability were needed. Bulk FHIR protocols needed to be expanded to match relevant records in other institutions. Fortunately, the HAPI FHIR standard has a master data management module (MDM) that can be called to merge records[15].

MDM algorithms in the HAPI server replaced the master person index in prior versions and offer a flexible approach to define matching algorithms [16]. There are many match algorithms that can be used. For example, the following will perform a Levenschtein distance string comparison on the first official given name. The match threshold identifies how close the two stings must be to result in a positive match on that demographic field.

~~~
{
 “name”: “firstname-leven”,
 “resourceType”: “Patient”,
 “fhirPath”: “name[0].where(use=‘official’ or use=‘usual’ or use.exists().not()).given[0]”,
 “similarity”: {
 “algorithm”: “LEVENSCHTEIN”,
 “matchThreshold”: 0.8
}
~~~

MDM performs asynchronous matching of patients as they are created or updated in the FHIR server. The results of the match determine if the patient is a match or not. When a match exists, the patient is linked to a Golden Resource that has a unique enterprise identifier (EID).

In order to expand retrievals to all sufficiently similar patients, HAPI FHIR was extended to include an _mdm flag for use in retrieval operations. In the BULK FHIR export, if the _mdm flag is set to true, the Golden Resources are used to return all patients matched to the supplied patients. When the Group resource, along with the _mdm=true flag, is passed in a Bulk FHIR request, the group of patients, as well as all patients matched in the MDM are used to return the set of resulting resources. The client application is not responsible for maintaining the MDM matching, but they are responsible for maintaining the Group status of their patients. The information returned will include the MDM EID allowing the results to be correlated. Without the _mdm flag, the client would need to query MDM for each patient to return the list of patient identifiers matched to their patient. They then would have to use that expanded list of identifiers in subsequent queries. With the _mdm flag, matching is always taken into account. This significantly reduces the implementation burden for population health systems.

The need to use MDM to perform matching also influences Use Cases (1b) and (2). In these use cases, there is no prior record of the patient in FHIR server, or the record is insufficiently detailed to allow accurate matching. In this setting, Group functionality becomes particularly important. Part of creating a group must be ensuring a sufficiently detailed record for matching exists at the time of the query using MDM functionality. This requires either updating or creating a record in the FHIR server for the patient, with sufficient demographic data and the institution’s patient identifier, for use by the FHIR server’s MDM to merge records as the group is defined. Creating a group needs to be a coordinated effort between the population health application and a FHIR server.

After testing this system in an internal sandbox, with 100’s of patients, we worked on scaling the system to tens of thousands of vaccine messages (V2.X) and thousands of simulated test cases in the American Immunization Registry Association (AIRA) IIS Sandbox. No further issues were identified requiring the customization of the FHIR server or other extensions to Bulk FHIR protocols.

### Population Health Tool

A group of 57 MUSC medical students successfully used the population health tool to conduct an outreach effort to the 1500 targeted patients. Up to 3 phone call attempts were made by the students to contact each patient totaling 2867 calls. 374 patients were reached (24.9%) and 225 patients declined a conversation (15.0%). 264 patients had an incorrect phone number listed in the EHR (17.6%).

The one-by-one query tool periodically failed to update records, with queries timing out or failing during peak periods of the pandemic. Failures occurred at all three postulated points: timing out of a query request on the REDCap side, timing out of a session on the IIS side, and case specification errors. Table 1 shows the errors observed at the beginning of outreach efforts (July 2021) and near the end (July 2021). Queries decreased as patients received vaccines or opted out of further contact.

**Table 1.** Types of errors observed with the one-by-one method over two one-month periods in the pandemic.

| Tracking period | 07/21 | 07/22 |
| --- | --- | --- |
| Total queries by VACtrac | 48,050 | 30,037 |
| Server time out errors | 813 | 5 |
| Session time out errors | 121 | 1 |
| Case specification error | 1729 | 220 |
| Average query response time | 932 ms | 391 ms |
| Percent queries failing | 5.54% | 0.75% |

## Discussion

Access to data from immunization registries for population health activities is a topic of increasing importance and a focus of the Center for Disease Control and Prevention’s new HELIOS initiative.[17] This application describes initial work to extend the Bulk FHIR protocol to address population vaccine management performed prior to the Helios initiative. This study may help inform those collaborative efforts in the HELIOS FHIR Accelerator.

As was quickly learned in agile development cycles, the most significant issue with the processing of a bulk query within a FHIR server is patient matching. Vaccination data can come from a variety of providers. Most IIS systems have some form of master-person-index solution.[4] However, a provider organization seeking access to IIS data may not have access to the IIS patient identifier, creating the need for matching algorithms embedded in the bulk query algorithm. The need for patient matching resulted in a requirement for sufficient demographic data to implement probabilistic algorithms. Our work found that ensuring such data exists in a patient record in the FHIR server needs to be accomplished prior to the submission of bulk query to achieve appropriate return rates to the query.

The question of the benefits of Bulk FHIR queries for population health purposes is still open. Using a FHIR server or another computational resource for mass querying seems likely to address limited capacity issues. The primary benefit of a FHIR server and a bulk query method is to allow asynchronous query and reporting, which further preserves limited computational resources for interactive queries for use within the context of one-on-one patient care and for the application of vaccine forecasting algorithms.

The availability of tools that use Bulk FHIR protocols to query patient data is also limited. Therefore, we developed a REDCap instance to manage bulk queries. This is, to our knowledge, the first implementation of a Bulk FHIR query tool for vaccine data. Student volunteers could use this program to manage an extensive vaccination outreach program. However, they could only connect to the state system in a one-at-time fashion to monitor the 1500 persons. The one-at-a-time implementation, initially had a high failure rate (5%), resulting in the data on vaccination status often not being updated. Given the adequate prior performance of the one-by-one approach in test beds and during periods of presumptive lower use, these results highlight the need for asynchronous bulk query capacity in IISs to respond to population health queries.

### Limitations

This study did not thoroughly test the Bulk FHIR protocol in a live clinical application. Instead, it was tested among several testbeds and in combination with a population health management tool that could use FHIR-derived protocols to query in bulk or individually. Many extensions to existing Bulk FHIR protocols were applicable, the discovery of which, along with the updating of the HAPI FHIR application with the necessary changes, being the point of the exercise. Ultimately the acceptability of advanced tools such as FHIR servers to IIS systems will depend on funding to states to acquire such systems and the availability of skilled personnel to use and maintain them.

## Conclusion

With extensions, and with integration into population health management tools, Bulk FHIR may be an important tool to help providers work with IIS data to address vaccination gaps.

## Data Availability

All data in the present study are available upon reasonable request to the authors.

## Acknowledgement

We would like to thank the American Immunization Registry Association (AIRA) and particularly Nathan Bunker and Mary Beth Kurilo for helpful discussions and access to and help with the use of the AIRA IIS test bed for testing of the VACTrac system.

## Software Availability

The HAPI FHIR server is available at http://hapifhir.io. The REDCap master file and middleware extensions for REDcap for Bulk FHIR and for one-by-one IIS query are available at (https://github.com/vactraccovid/vactrac).

## Notes

### Competing Interest Statement

Two authors (JA and DW) are members of a commercial enterprise, Smile Digital Health, which sells an enhanced, supported version of the open-source HAPI FHIR server.

### Funding Statement

This publication was supported, in part, by the National Center for Advancing Translational Sciences of the National Institutes of Health under Grant Number UL1 TR001450. The content is solely the responsibility of the authors and does not necessarily represent the official views of the National Institutes of Health.

### Author Declarations

This work was classified as a quality improvement activity by the Institutional Review Board of the Medical University of South Carolina.

